# Deep representation learning of electrocardiogram reveals novel insights in cardiac structure and functions and connections to cardiovascular diseases

**DOI:** 10.1101/2023.12.05.23299459

**Authors:** Ming Wai Yeung, Rutger R van de Leur, Jan Walter Benjamins, Melle B. Vessies, Bram Ruijsink, Esther Puyol-Antón, J. Peter van Tintelen, Niek Verweij, Rene van Es, Pim van der Harst

## Abstract

**Background:** Conventional approaches to analysing electrocardiograms (ECG) in fragmented parameters (such as the PR interval) ignored the high dimensionality of data which might result in omission of subtle information content relevant the cardiac biology. Deep representation learning of ECG may reveal novel insights.

**Methods:** We finetuned an unsupervised variational auto-encoder (VAE), originally trained on over 1.1 million 12-lead ECG, to learn the underlying distributions of the median beat ECG morphology of 41,927 UK Biobank participants. We explored the relationship between the latent representations (latent factors) and traditional ECG parameters, cardiac magnetic resonance (CMR)-derived structural and functional phenotypes. We assessed the association of the latent factors with various cardiac and cardiometabolic diseases and further investigated their predictive value for cardiovascular mortality. Finally, we studied genetic components of the latent factors by genome wide association study (GWAS).

**Results:** The latent factors showed differential correlation patterns with conventional ECG parameters with the highest correlations observed in factor 8 and PR interval (r=0.76). Multivariable analyses of the ECG latent factors recapitulated CMR-derived parameters with a better performance for the left ventricle than the right. We saw higher performance in models for structural parameters than functional parameters and observed the highest adjusted R^2^ of 0.488 for left ventricular LV end-diastolic mass (LVEDM). The latent factors showed strong association with cardiac diseases. This included bundle branch block and latent factor 28 (OR= 2.72 [95% confidence interval CI,2.46-3.01] per standard deviation, SD change); per SD change of latent factor 27 was associated with cardiomyopathy (OR=2.38, 95%CI 1.97-2.89) and heart failure (OR=1.94, 95%CI 1.71-2.21). In the GWAS of the latent factors, we identified 170 genetic loci with 29 not previously associated with electrocardiographic traits. Following up with bioinformatic analyses, we found the genetic signals involved in cardiac development, contractility and electrophysiology.

**Conclusions:** Deep representation learning of 12-lead ECG provided not only clinically meaningful but also novel insights into cardiac biology and cardiovascular health.

Graphical abstract

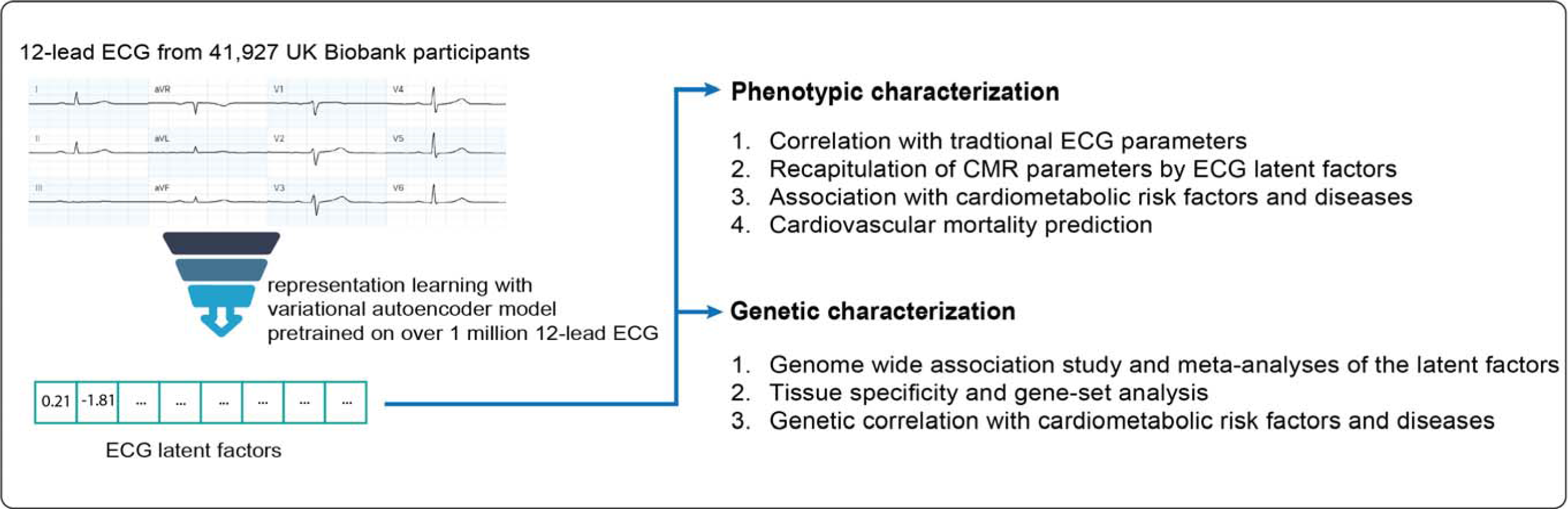

## Introduction

The electrocardiogram (ECG) is a non-invasive test that registers the electrical activity of the heart muscle cells (cardiomyocytes) allowing the evaluation of its function and detection of disorders. Clinical interpretation of the ECG is a pattern comparison task anchored to predefined features, namely the waveforms (such as P-wave and T wave) and intervals (the QRS complex). To date, large scale genome wide association studies of these predefined features have yielded hundreds of genetic loci linked to cardiac electric functions^1–3^. While these well-defined features are clinically robust, they may not fully capture the information content contained in the ECG as there may exist morphologies informative to the cardiac electrophysiology not measured in such features or even obscure to human eyes. To this end, we demonstrated in a previous study that a high-dimensional analysis of the complete ECG provided additional biological insights^4^.

Variational autoencoder (VAE) is an artificial neural network architecture for unsupervised representation learning^5^. While VAE is similar architecturally to a classical autoencoder (both with an encoder which maps input to latent representation and a decoder component to map the latent space to input), VAE learns the underlying probability distributions of the data. We hypothesized that VAE could be utilized for uncovering additional insights from the ECG beyond traditional descriptors and that genetic study of the latent representation of the ECG would expand our knowledge of genetic factors linked to the cardiac conduction system. In the current study, we employed β-VAE^6^, a variant of VAE for improved disentanglement of the latent representation, to learn the disentangled latent representation of the 12-lead ECG. We finetuned a β-VAE model, pretrained on 1,144,331 12-lead ECGs^7^, to generate 32 latent factors from the 12-lead ECG of 40,185 UK Biobank participants. We investigated the associations between these disentangled ECG factors and structural and functional parameters of the heart derived from cardiac magnetic resonance (CMR) as well as cardiovascular risk factors and diseases. We then performed genome wide association studies on these factors with the aim to identify additional genetic variants associated with ECG morphologies. To inform the biological context, we further characterize our genetic findings by gene-set clustering analyses. Finally, we exploited the decoder component of the β-VAE model to visualize the genetics associations to the ECG morphologies.

## Methods

### Study Population

The UK Biobank study is a population-based prospective cohort in the United Kingdom in which approximately 500,000 individuals aged between 40 and 69 years were included between 2006-2010. All participants have given informed consent for this study. The UK Biobank has ethical approval from North West - Haydock Research Ethics Committee (REC reference: 16/NW/0274). Details of the UK Biobank study has been described in detail previously^8^. This research has been conducted using the UK Biobank Resource under Application Number 74395. Disease outcomes were captured based on a composite source of data from interview with a trained nurse at the visit to assessment centres (self-report) and linked electronic health records including hospital inpatient episode data and primary care data processed using the R package ukbpheno^9^. The last follow-up dates were 30 September 2021 for English participants, 31 July 2021 for Scottish participants and 28 February 2018 for Welsh participants.

### ECG processing

A subset of the UK Biobank participants was invited for the imaging visit. During the imaging visit, each participant received a 10-second 12-lead ECG measurement at rest in a lying position. The ECGs were acquired at 500Hz and converted to median beats using the GE Cardiosoft system and were recorded in XML format (Data-Field 20205 in the UK Biobank resource). Traditional ECG descriptors including ventricular rate, P interval, QRS duration, QT interval, P axis, R axis and T axis were derived for each ECG using the GE system.

### CMR-derived structural and functional parameters

CMR was acquired at the same visit during which the 12-lead ECG was acquired. Short-axis and 2- and 4-chamber long-axis cine CMR images were acquired using a clinical wide bore 1.5 Tesla scanner (MAGNETOM Aera, Syngo Platform VD13A, Siemens Healthcare, Erlangen, Germany) in the absence of a pharmacological stressor or contrast agent^10^. We applied a validated DL-based pipeline to derive the structural and functional parameters of the heart from the CMR images^11^. The structural parameters including end-systolic volume (ESV), stroke volume (SV) of the left ventricle (LV) and right ventricle (RV) respectively as well as LV end-diastolic mass (EDM) and mean global myocardial wall thickness were estimated. We indexed the volume and mass estimates by body surface area (BSA) to correct for overall body size and we additionally computed LV mass-to-volume ratio by dividing LVEDM by LVESV. Functional parameters including LV / RV peak ejection rate, peak filling rate and ejection fraction as well as LV mitral annular plane systolic excursion (MAPSE) and RV tricuspid annular plane systolic excursion (TAPSE) were estimated. In addition to the quality control steps of the DL-based pipeline^11^, we removed outliers which were defined as values 3 interquartile ranges below the first or above the third quartile in the current study.

### Unsupervised representation learning of ECG

We applied an artificial neural network β-VAE to learn factors from the median beat ECG in an unsupervised manner. Briefly, a VAE is a generative model with an encoder-decoder architecture. The encoder transforms input, here the original 12-lead median beat ECG, to the latent factors which follow Gaussian distributions, whereas the latent factors could be mapped back to the input space by the decoder resulting in a reconstructed ECG. β-VAE is a VAE variant that is encouraged to learn factorised latent factors through weighing (β-value) of the loss term during the training process. As a result, β-VAE models tend to produce better disentangled (statistically orthogonal) latent representation following the multivariate Gaussian distributions.

In the current study, we finetuned a β-VAE model with a latent space of 32 dimensions pretrained on 1,144,331 ECGs recorded on a GE MAC 5500 (GE Healthcare, Chicago, IL) from 251,473 patients primarily of Dutch origins^7^ on the ECGs of 41,927 UK Biobank participants. The median beat ECGs are computed by the GE system were resampled to 500Hz and a 1^st^ order polynomial was substracted to remove baseline wander. During training, a weight of 10 was applied to the samples in the QRS complex. Data was split in a 90:10 manner and the epoch with the lowest loss in the validation dataset was chosen. To validate the quality of the reconstructions, we calculated a Pearson correlation coefficient for the orginal and reconstructed median beat ECGs. All ECGs with a correlation below 0.7 were excluded. The standardized latent factors from the finetuned model were taken forward for downstream analyses. A detailed description of the model architecture and loss can be found in a previous study.^7^

The influence of the individual ECG factor on the median beat ECG morphology was visualized using factor traversals. These were derived by varying the values of the factors individually between −3 and 3, while generating the median beat ECG using the decoder. As the other factors are kept constant, the individual influence of that factor on the ECG morphology can be visualized.

### Ascertainment of health outcomes

We interrogated the associations of the ECG latent factors with various cardiovascular, metabolic as well as rhythm disorders listed in the **Table S1**. Ascertainment of these diseases in the UK Biobank was based on a combination of hospital inpatient records, primary care records and self-reported records during an interview with a trained staff. Mortality including cause (coded by ICD-10) and date of death were captured from death registries^12^. Definitions of the health outcomes are presented in (**Table S1**). Participant follow-up started at the date of imaging visit (where the 12-lead ECG and CMR were performed) and ended at date of event, death or last recorded follow-up, whichever occurred first. Participant follow-up was censored on September 30, 2021 for participants from England / Wales and October 30, 2021 for participants from Scotland respectively. Health outcome data was processed and extracted using the ukbpheno v1.0 package in R^9^.

### Phenotypic characterization of the latent factors

We first assessed the Pearson correlations between the latent factors and traditional ECG descriptors in order to explore the information captured by the factors. We further investigated whether cardiac structure and functions could be inferred from the latent factors derived from the 12-lead ECG. More specifically, we regressed each CMR-derived structural and functional parameter on all 32 latent factors. Finally, we performed association analyses with health outcomes using logistic regression as well as mortality (all-cause mortality and cardiovascular mortality) prediction using Cox proportional hazards models. We included age and sex as covariates in all regression models.

### Genome wide association study and candidate genes

We conducted genome wide association study (GWAS) on participants with genotyping data available. Two custom Affymetrix Axiom^TM^ (UK Biobank Lung Exome Variant Evaluation or UK Biobank) genotyping arrays with >95% common content were used. Imputation was performed using Haplotype Reference Consortium as the primary reference panel with addition of merged UK10K and 1000 Genome phase 3 reference panels. Quality control of samples and variants, as well as of the imputation was performed by the Wellcome Trust Centre for Human Genetics^8^. The current study was performed on the imputed data supplied by the UK Biobank. Individuals with overall missingness >5% or excessive heterozygosity, and individuals whose genetically inferred sex did not match with the reported sex, were excluded from the analyses. Additionally, variants with a minor allele frequency smaller than 0.5% or an INFO-score smaller than 0.3 were excluded.

We first performed GWAS of autosomes for individual latent factors using BOLT-LMM v2.3.1^13^. A linear mixed model was fitted for each of the z-transformed latent representation. GWAS analyses were adjusted for age at the time of the imaging visit, sex, genotyping array and the first 30 principal components (provided by UK Biobank) to adjust for population stratification. Individuals with missing information on any of the covariates were excluded from the GWAS analyses. The nearest protein coding gene and any additional gene within 10kb of each of all independent lead variant (with *P*<5×10^-8^ and with LD r^2^>0.005) were annotated as candidate causal genes. We defined the region ±500KB around the independent lead variant as a genetic locus. To identify novel associations, we cross-referenced our findings with 7,464 significant associations for electrocardiography (EFO_0004327) including all child traits in the GWAS catalog (record downloaded on 27-06-2023). To take into account of multiple testing, we estimated the number of effective independent tests for the 32 latent factors using an eigenvalue ratio measure^14,15^. With 23 effective independent tests, a more stringent p-value of 2.17×10^-9^ (5×10^-8^/23) was taken as Bonferroni corrected genome wide significance.

To further refine the signals detected in GWAS, we applied statistical fine-mapping on all loci identified to prioritize putative causal variants. Bayesian fine-mapping was performed on summary statistics using FINEMAP (v1.4)^16^. A shotgun stochastic search method was used to produce 95% credible sets under the assumption of a number of causal variants (*k)* from 1 up to 5, each with estimated posterior probabilities which jointly summed up to 1. We considered the variants in the top causal configuration under *k* with the highest posterior probabilities as the likely causal variants. For each likely causal variant, we searched for coding variants in high LD (R^2^C>C0.8) with dbNSFP (v.4.2)^17^.

Genetic correlations between the latent factors were estimate using LD score regression^18^. Given the higher genetic correlations observed in factors with small phenotypic variance, we performed meta-analyses on all factors with significant genetic correlations r_g_>0.4 after Bonferroni correction (p<0.05/496=1.01 ×10^-4^) using Multi-Trait Analysis of GWAS (MTAG)^19^ with the objective of identifying additional genetic signals embedded in the ECG. MTAG is a tool for analysis of multiple GWAS summary statistics which applies a generalized inverse-variance-weighted meta-analysis that allows for sample overlap. In MTAG analyses we considered common variants with minor allele frequency larger than 0.01 and INFO-score larger than 0.3.

### Tissue specificity and gene-set analysis

To further interpret the genetic composition of the latent factors, we applied Multi-marker Analysis for GenoMic Annotation (MAGMA) tool for gene property analysis and gene-set analysis^20^. MAGMA analysis was done via the platform Functional Mapping and Annotation of Genome-Wide Association Studies (FUMA) v1.4.0 with MAGMA v1.08, using default settings. Briefly, gene-based test statistics were first obtained with a SNP-wide mean model which were then converted to Z scores. To identify tissue specificity of the GWAS associations, these gene-based Z scores was submitted to gene property analysis in which the Z scores were regressed against expression data set GTEx v8 which including 52 tissues and two cell lines across 30 general tissue types.

Enrichment of association was tested with 15,485 predefined gene-sets from MsigDB v7.0^21^. The database contains 5,497 curated gene sets from 9 pathway databases including BioCarta, Kyoto encyclopaedia of gene (KEGG), Reactome, WikiPathways, Matrisome Project, Pathway Interaction Database, SigmaAldrich database, Signaling Gateway database and SuperArray SABiosciences database, as well as 9,988 Gene Ontology (GO) terms organized in three categories, namely biological processes (BP), cellular components (CC) and molecular functions (MF). Gene-sets that reached statistical significance after Bonferroni correction (*P=* 3.23×10^-6^) were then put forward for clustering analysis using R package simplifyEnrichment (v1.1.5)^22^. Semantic similarity between GO terms per categories, namely BP, MF and CC, were measured by relative relevance^23^ while similarity between non-GO curated gene-sets were measured by kappa coefficients^24^. Clustering of the gene sets was performed using the binary cut algorithm, which recursively divides terms into 2 groups by medoids^22^. Based on the resulting functional clusters, we visualized the enriched gene-sets in Voronoi treemaps^25^. Voronoi treemap is a variant of treemap consists of polygonal cells instead of rectangular ones. We set the size of the cell to be proportional to the binary logarithm of the inverse of the p-value in MAGMA association test while darker shade of the cell reflects larger relative effect size of the gene-set within the cluster. We generated Voronoi treemaps for all latent representation with significant gene-sets using the R package WeightedTreemaps.

### Morphological manifestations of the genetic components and associations with clinical outcomes

To visualize morphological effects of the genetic variants, we modified each latent representation by the effect size associated for the target variant in GWAS and then reconstruct the ECG by submitting the modified latent factors the decoder of the VAE model. More specifically, we transformed all effect sizes of these associations back to original scale and used them to modify the average latent factors in an additive manner. We considered associations with suggestive p-value <1×10^-5^ only to reduce noise by potentially spurious associations in this visualization task. We made an interactive tool to visualize the morphological effects of the genetic variants which is available online via https://genetics.ecgx.ai/.

We estimated genetic correlations between the latent factors and a range cardiometabolic as well as rhythm disorders (**Table S2**) in order to investigate the shared genetic components using linkage disequilibrium score regression (LDSC) v1.0.1^18^. Genetic associations of the diseases were done within the subset of unrelated UK Biobank participants without 12-lead rest ECG. To obtain reliable genetic correlation estimates, we included the phenotypes if all three criteria were met in the SNP heritability analysis: the Z-score is 1.5 or above, the mean Chi-square of the test statistics is larger than 1.02 and the intercept is between 0.9 and 1.1^26^.

## Results

### Clinical characteristics and AI-derived phenotypes

Out of 502,408 UK Biobank participants, 12-lead ECG data was available for 41,927 participants in the current analysis. Clinical characteristics of these participants are shown in **Table S3**. The mean age of these participants at the time of visit was 64.6 (standard deviation 7.73 years) and 51.6% were female. We took the median beat of ECG passing quality control (QC) from 41,581 UK Biobank participants to finetune the β-VAE model originally trained on 1,144,331 ECGs from 251C473 patients. Reconstruction of the ECG using the finetuned model achieved a Pearson correlation of 0.963. We then transformed the ECG into 32 latent factors using the encoder of the trained model for downstream analyses. Kernel density estimate plots of each latent factors over age by sex can be found in **Figure S1. Factor traversals for visualizing the effect of individual ECG factors on the ECG morphology can be found in Figure S2.** Disentanglement was successful between 21 latent factors with variance close to one while higher phenotypic correlations were observed for the 11 latent factors of subtle morphologies (with variance <0.001), namely factor 2, factor 3, factor 4, factor 7, factor 14, factor 18, factor 20, factor 21, factor 24, factor 28 and factor 29 (**Figure S3 and Table S4).**

CMR phenotypes derived from the respective AI-based pipeline were available for 34,021 participants who also had latent factors from ECG available. **Table S5** reports the summary statistics of these phenotypes by sex.

### Association between latent factors and traditional ECG parameters

To demonstrate that the latent factors also capture electrophysiological information contained the traditional ECG parameters, we performed Pearson correlation analyses between the latent factors and the traditional ECG parameters (**Figure 1**). The strongest correlation was observed between factor 10 and ventricular rate (r=0.71). PR interval was strongly correlated with factor 8 (r=0.76) while QRS was most correlated with factor 25 (r=--.66) and QT interval with factor 30 (r= −0.7). Other strong correlations were observed for factor 31 with R-axis (r= −0.52) and T-axis (−0.53) respectively.

**Figure 1.**
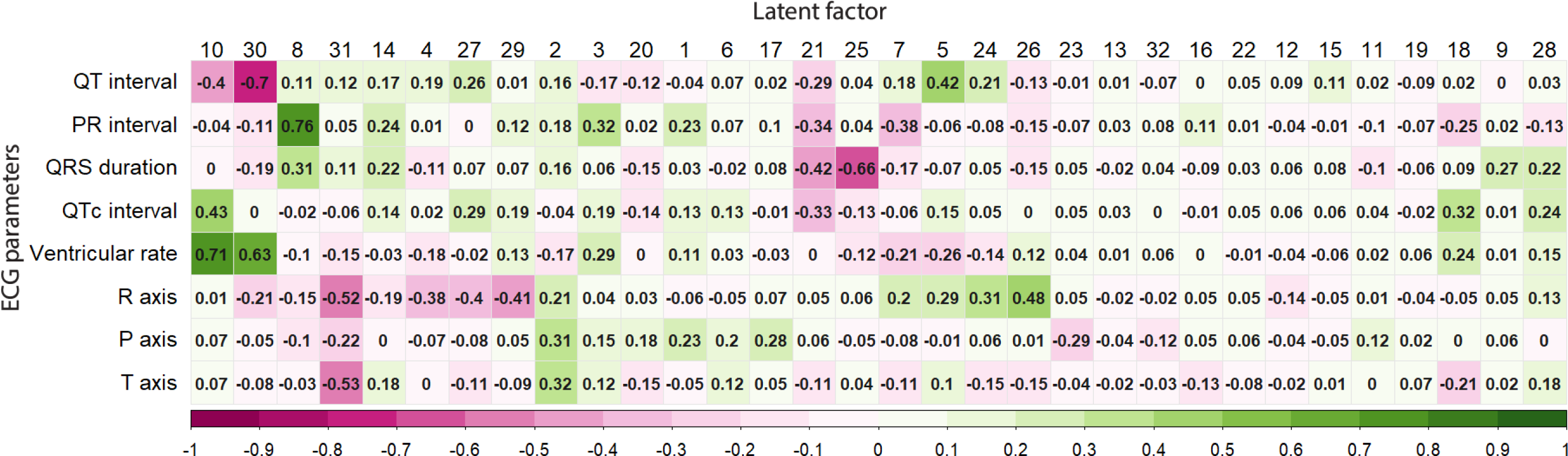
ECG latent factors capture information represented in traditional ECG parameters. Pearson correlation coefficients between electrocardiogram measures of PR interval, QRS duration, QT interval, Bazett corrected QT (QTc) interval, ventricular rate, P-axis, R-axis, and T-axis, and latent factors generated from the 12-lead ECG of 41,927 UK Biobank participants.

### Recapitulation of CMR parameters by ECG latent factors

We built multivariable linear regression models of each CMR parameter againset age at imaging, sex and all 32 latent factors and assessed the goodness of fit using the adjusted R^2^. The variations accounted for in the models for structural CMR parameters, namely LVEDM (adjusted R^2^ = 0.488) and LV mean global myocardial wall thickness (adjusted R^2^ = 0.470) were higher, as compared with functional parameters (**Figure 2)**. Considering the effect sizes, factor 32 and 18 were the strongest predictors positively associated with both structural parameters while factor 29 and 26 were negatively associated. Consistently factor 26 and 32 were also strongly associated with LV functional parameters including LVSV, LVPER and LVPFR. Associations of RV parameters with ECG latent factors were weaker compared with LV counterparts (**Figure 2)**. Latent factors 16, 11 and 15 were strongly associated with RV functions. Notably, factor 30 was negatively associated with both stroke volume estimates while factor 20 was positively associated with both LV/RV ejection fraction estimates. It was worth to note that LVEF, an important clinical measure of LV systolic function, as well as its surrogate measure MAPSE were only weakly explained by the ECG latent factors with an adjusted R^2^ of 0.14 and 0.069 respectively.

**Figure 2.**
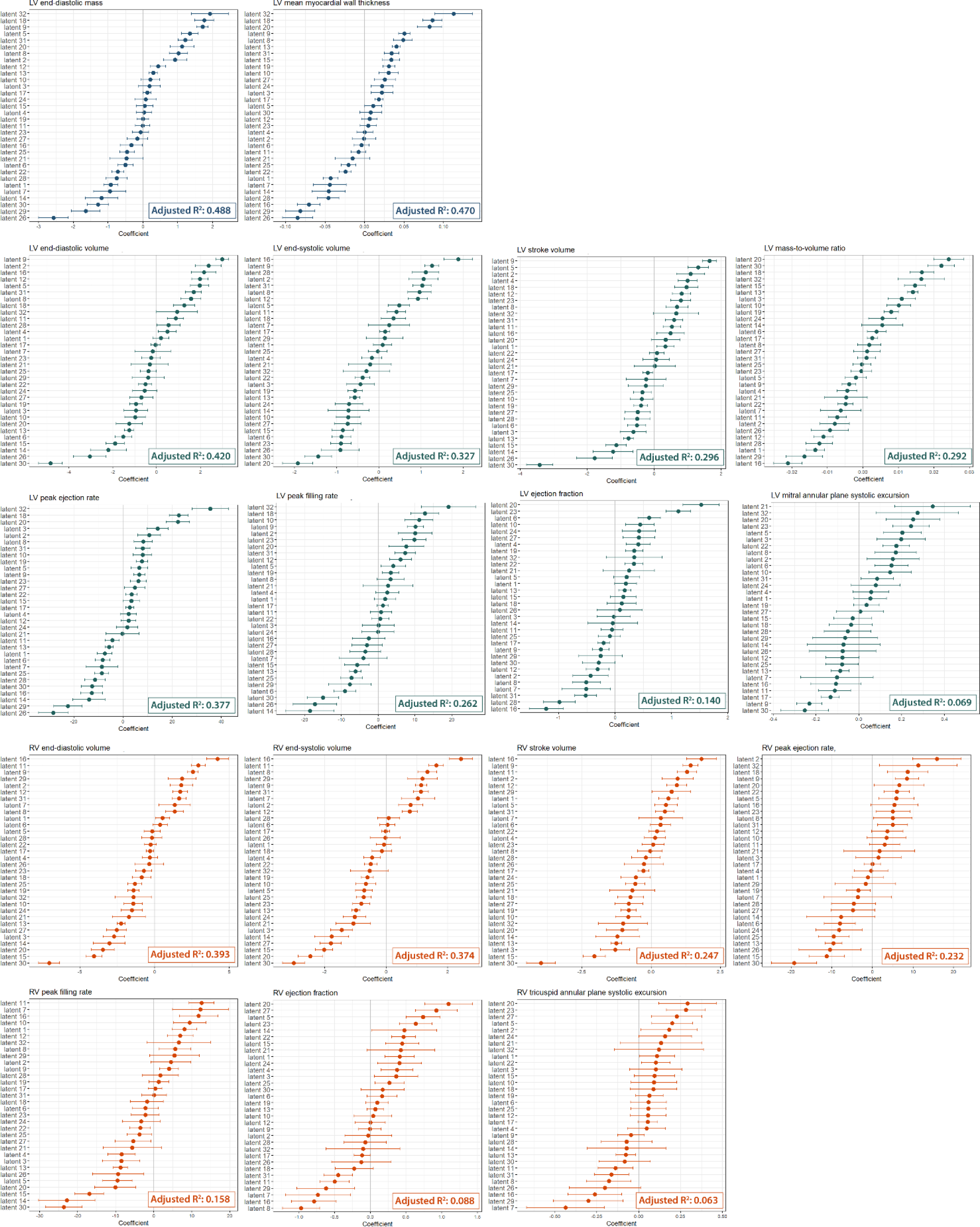
Association of ECG latent factors with CMR parameters. Regression coefficients of the latent factors in the multivariate linear regression on each CMR-derived structural and functional parameter. The coefficient describes the effect per standard deviation change of the latent factor. Adjusted R^2^ indicates the goodness-of-fit of the linear regression model.

### Association with cardiometabolic outcomes

We then explored the relationship between latent factors and selected health outcomes. **Table S6** reports the significant associations of each factor for the health outcomes in a logistic regression model adjusted for age and sex. We found the strongest associations for conduction disorder bundle branch block but also strong association for rhythm disorders like atrial fibrillation as well as structural heart disease like cardiomyopathy. Compared with the conduction and rhythmic disorders in which information was captured by few specific latent factors as two distinct clusters (**Figure 3**), we observed more moderate but scattered associations for cardiometabolic diseases such as hypertension, coronary artery disease and hyperlipidemia.

**Figure 3.**
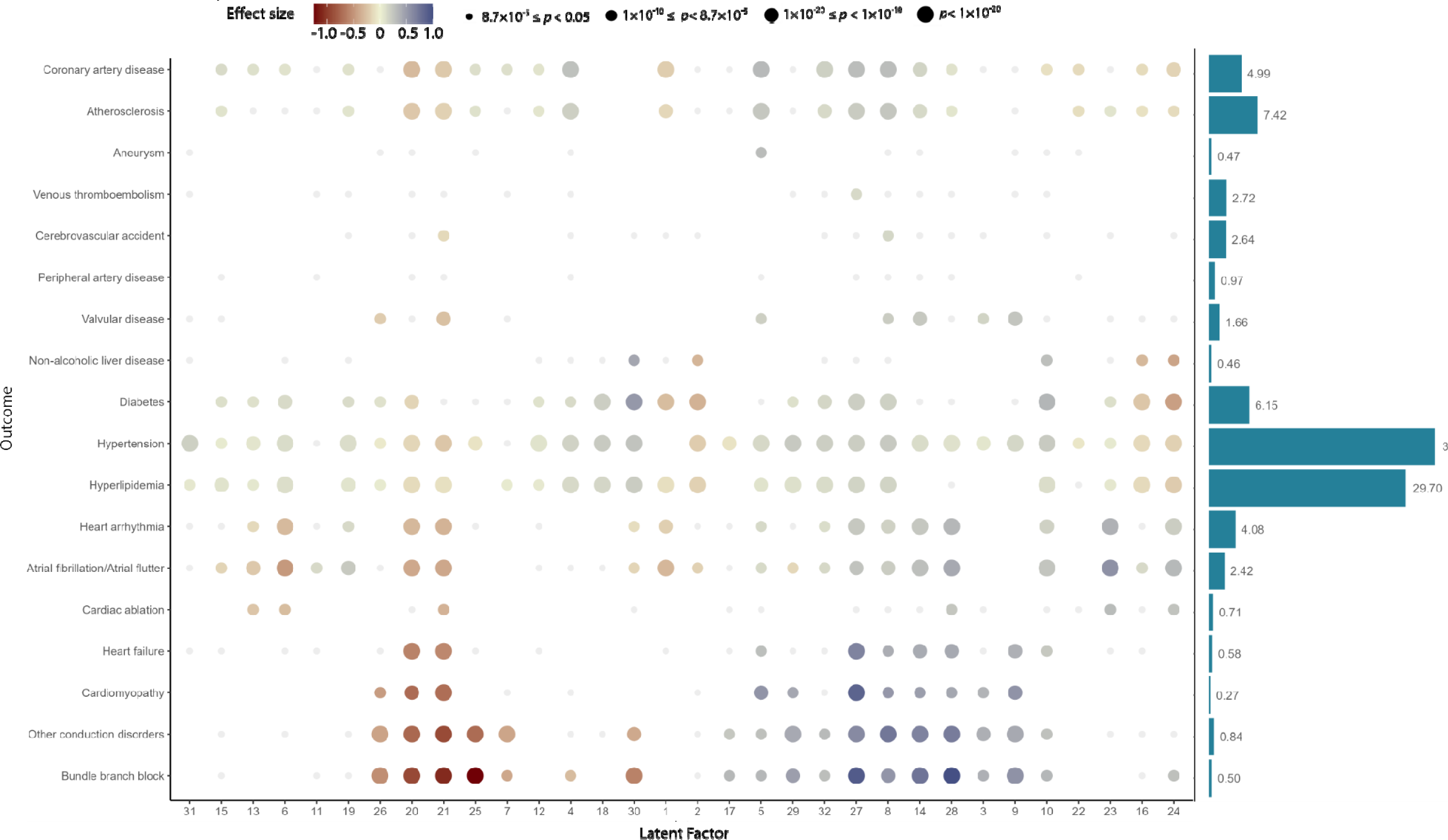
Associations of latent factors with cardiometabolic diseases. Dot plot shows the association of the latent factors as a risk factor for selected cardiometabolic disease. The colour of the dot indicates the coefficients in logistic regression model. Coefficients were adjusted for age and sex. The size of dot reflects the p-value categories for the latent factor in the correspond logistic regression model; p=8.7 ×10^-5^ is the Bonferroni corrected p-value taking α=0.05. The bar plot shows the prevalence of the disease at the imaging v when the ECG was acquired.

### CVD mortality prediction

During the median follow-up of 3.4 (interquartile range 2.5 - 4.9) years, 187 participants died of CVD. Six out of 32 latent factors, namely factor 8, 9, 10, 14, 20, 21 and 28 were associated with CVD mortality with *p*<0.002 (**Figure S4**). After adjustment for age and sex, factor 8 and factor 10 were the strongest predictors positively associated with mortality with hazard ratio, HR of 1.35 (95% confidence interval, CI 1.17 - 1.56) and 1.34 (95% CI 1.18 - 1.53) respectively; this was contrasted by factor 20 and 21 which were negatively associated with mortality with a HR of 0.75 (95% CI 0.65 - 0.87) and 0.76 (95% CI 0.67 - 0.86) respectively. For comparison we tested the association with z-transformed values of the traditional ECG parameters (**Figure S5**). Ventricular rate was the strongest predictor with a HR of 1.28 (95% CI 1.13 – 1.45) and this was followed by corrected QT interval with HR of 1.25 (95% CI 1.14 – 1.36) and QRS duration with HR of 1.22 (95% CI 1.10 – 1.36).

### GWAS and MTAG analysis

Genetic data were available for 40,815 participants at the time of analysis. We identified 164 independent lead variants at p<5×10^-8^, 104 of which with p<2.17×10^-9^, in the GWAS on the 32 latent factors (**Figure 4A and Table S7**). Notably 73 of them were identified in the 11 latent factors of subtle morphologies (**Table S4**). Comparison with records in the GWAS catalog revealed that the majority of the loci, namely 141 loci, in the current GWAS had been reported for ECG morphologies in previous studies (**Table S7)**. Among 23 novel loci with lead variants reaching p < 5× 10^-8^, six (nearest coding genes *EFEMP1, ERBB4, MAP9, SNCAIP* and *GPR126*) passed the more stringent p<2.17×10^-9^. Regional association plots for the novel loci are presented in **Figure S6.** We visualized the effects of lead genetic variants on the ECG morphologies via the decoder model of the VAE which is available at https://genetics.ecgx.ai/ (**Figure S7**).

**Figure 4.**
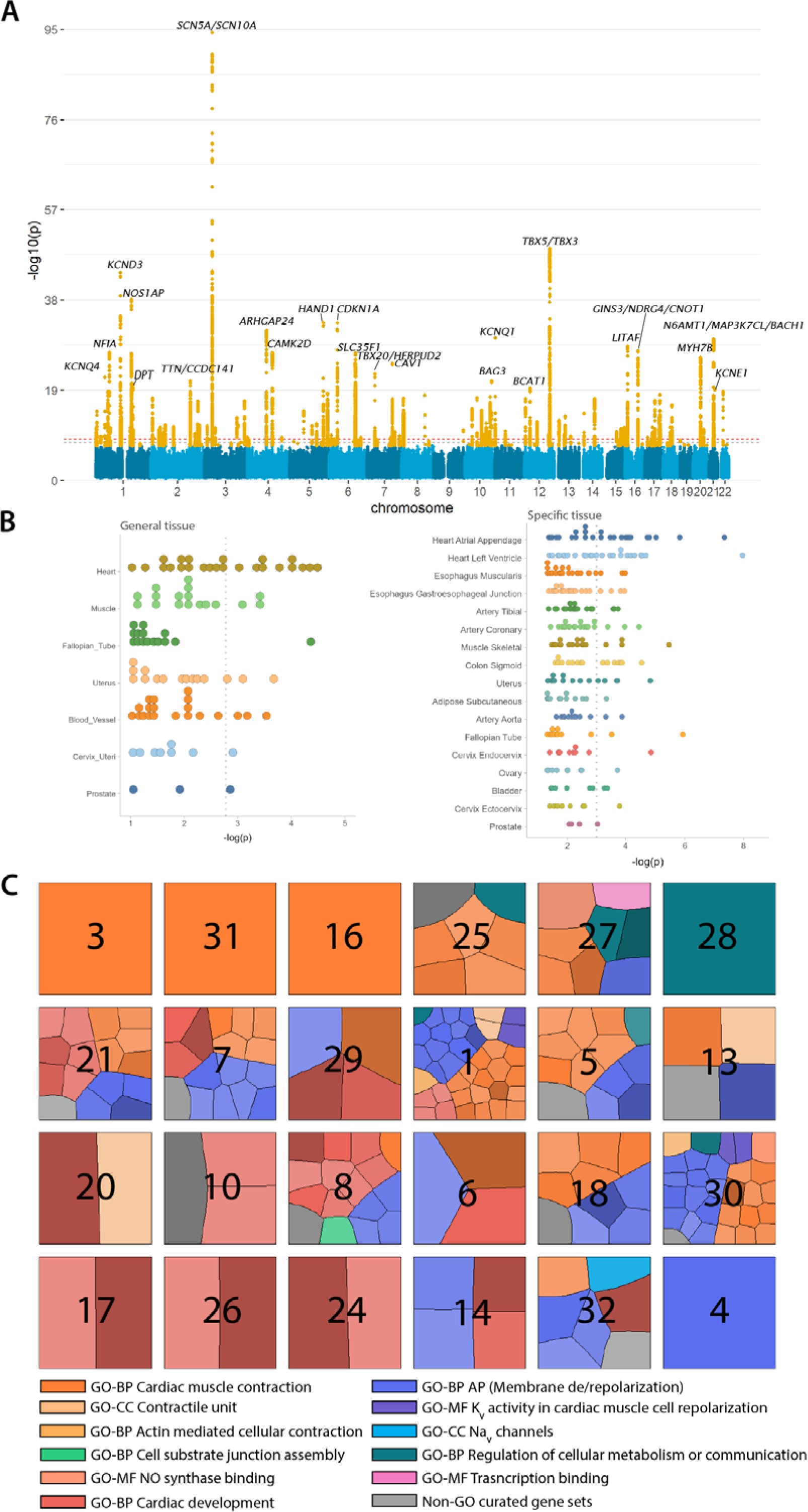
Enrichment in cardiac tissue and pathways for genetic variants identified in genome wide association study of the latent factors. (A) Manhattan plot of all latent factor (smallest p-value across latent factors is shown); variants in orange indicate those pass p<5×10^-8^ and the top loci have been annotated with their nearby genes. (B) Tissue specificity of the latent factors with GTEx datasets (v8). Results with p<0.05 in MAGMA tissue expression analysis are shown. Each dot represents one latent factor and dotted line indicate the Bonferroni corrected p-value. Left: general tissue types; right: specific tissue types. (C) Clustering of the gene sets associated with each latent factor visualized in Voronoi tree maps. Clustering was performed on Gene Ontology (GO) associated gene-sets by semantic similarity and for non-GO curated gene-sets by kappa coefficients. Each cell within the tree map represents one gene-set with cell size reflecting the relative significance and darker shade reflecting stronger effect size among all gene sets for that latent in the MAGMA gene-set analysis.

Considering the significant genetic correlations for 11 latent factors with other latent factors (**Figure S3**), we performed joint analyses of these factors with genetically correlated factors (r_g_ ≥0.4) using MTAG^19^. We identified 6 additional novel loci close to *PTP4A2, HNRNPLL, ST5, GPC6, SELENOV and C20orf187* respectively at p<5×10^-8^ (**Table S8**). GWAS results including the joint analyses with novel loci can be explored online in https://my.locuszoom.org/.

### Candidate causal variants

To refine the signals identified in GWAS, we first performed statistical fine-mapping to prioritize candidate causal variants. The candidate causal configurations with the highest posterior probabilities for each of the loci are reported in **Table S7 and S8**. The lead variant remained the candidate causal signal selected by FINEMAP in most of the loci. Notably, the GWAS of factor 23 FINEMAP identified an additional signal rs2061770 (intronic of *SNX24*) at *SNCAIP* locus, a locus associated with heart rate response to exercise^27^. Two additional signals rs78656993 and rs1411924 were identified within *NRP1* gene from GWAS of factor 27; of note, rs78656993 is in an enhancer region (Ensembl regulatory feature ID: ENSR00000026629) while rs1411924 is located at a transcriptional factor binding site (ENSR00000400533). At the *DPP6* locus from GWAS of factor 3 an additional signal rs113687675 was identified and it is located at approximately 2,175 base pairs upstream of rs606231226, the variant reported for familial paroxysmal ventricular fibrillation in a Dutch family^28^. FINEMAP revealed n missense variant of *SEPTIN3* at the *NFAM1* locus from the GWAS of factor 28.

We next queried dbNSFP for all the candidate causal variants as well as their LD buddies (R^2^>0.8) to identify nonsynonymous coding variants (**Table S9)**. We found coding variants in known cardiomyopathies associated genes^29^ including *TNNT* (rs764862951), *TTN* (rs9808377, rs3829746, rs2042996, rs1560221, rs1001238, rs2042995, rs12693164, rs13390491, rs16866465 and rs12693166), *BAG3* (rs2234962), *JUP* (rs41283425) and *MYBPC3* (rs3729989) as well as in known rhythm disorders associated genes^30^ *KCNE1* (rs1805128). Among the novel loci identified in this current study with p < 5× 10^-8^, we observed missense variants for *AFAP1* (rs28406288), *ST5* (rs3794153), *SUGP1* (rs17751061) and *HAUS5* (rs71353000). Variant rs28406288 and rs17751061 had CADD scores above 20 (22.8 and 31 respectively) and were predicted to be pathogenic by several variant pathogenicity prediction tools including SIFT, Polyphen2 and Mutation Taster except for PrimateAI (**Table S9)**.

### Tissue specificity and gene-set analyses

Using the GTEx dataset, we observed the strongest enrichment in heart tissues in both general tissue types and specific types. (**Figure 4B, Table S10 and Table S11)**. A total of 11 latent factors were associated with heart tissue with p<0.0017 and 15 latent factors were associated with at least one of the heart specific tissue types arial appendage or left ventricle with p<9.6 ×10^-4^ after Bonferroni correction. Beside the heart, muscle and blood vessels were among the top enriched tissues with the strongest association observed for factor 24 (p = 5.18× 10^-6^) and factor 29 (p = 2.90× 10^-4^) respectively (**Table S11**).

To summarize the genetic signals associated with the latent factors into biological processes, we performed MAGMA gene-set enrichment analysis. A total of 191 gene-sets passed Bonferroni corrected significance threshold for 24 latent factors (**Table S12**). Latent factors with the most associations included factor 1 (n_gene_set_= 38) and factor 30 (n_gene_set_= 29), while 5 latent factors (factor 3, factor 4, factor 16, factor 28 and factor 31) were significantly associated with one gene-set. Clustering analysis of the gene-sets resulted in 12 functional groups (**Figure S8**) belonging to three main aspects of cardiac electrophysiology, namely cardiac development, muscle contraction and membrane de/repolarization (**Figure 4C**). Most latent factors were associated with gene-sets of multiple functional groups with some exceptions. Latent factors strongly enriched for development or tissue regeneration of the heart included factor 17, factor 24 and factor 26. This contrasted with factor 3, factor 16 and factor 31 which captured cell signalling of the His-Purkinje system.

### Shared genetic components with health outcomes

To explore the genetic overlap between the latent factors and health outcomes, we performed GWAS on the health outcomes in 446,014 UK Biobank participants who did not have 12-lead ECG recorded. We estimated the genetic correlations between 17 health outcomes and the latent factors with LD score regression. A total of 30 latent factors showed moderate genetic correlations with one or more cardiometabolic outcomes with *p*<0.05, among which 16 had correlations with a more stringent *p*<9.19×10^-5^ **(Figure S9 and Table S13**). The strongest genetic correlation was observed between factor 25 and bundle branch block *r_g_* = −0.69 (*P*=1.14×10^-9^). Factor 18 showed strong genetic correlation with bundle branch block as well, although in the opposite direction *r_g_* = 0.39 (*P*=3.43×10^-5^). We also observed strong genetic correlation for other conduction disorders with factor 8 *r_g_*= 0.46 (*P*=1.46×10^-10^), factor 25 *r_g_* = −0.42 (*P*=3.55×10^-7^) and factor 21 *r_g_* = −0.41 (*P*=1.82×10^-8^). In contrast, strong genetic correlations (|*r_g_* |>0.3) were observed for non-alcoholic liver disease with factor 30, factor 24 and factor 10; genetic correlations with vascular diseases including aneurysm, hypertension and peripheral artery disease was observed in factor 4.

On a locus level, we found 146 loci which have been previously reported covering 332 traits in studies documented in the GWAS catalog (**Table S14**). Besides electrocardiographic traits and rhythm disorders, which are most frequently reported, we observed great overlap in loci for physical measures including height (number of loci shared, n=34), Waist-to-hip ratio / waist circumference (n=19), blood pressure traits (n=32) and heel bone mineral density (n=10). Four of the novel loci, namely *MECOM*, *AFAP1*, *CCDC92/ZNF664/FAM101A* and *SNCAIP* have been associated with non-electrographic cardiac traits (**Figure S10**); the *HIST1H3G/ HIST1H2BI* locus has been associated with primarily lipid traits while the *MAP9* locus has been associated with vascular traits.

## Discussion

In this large study, we described the full 12-lead ECG waveforms in a latent space of 32 joint distributions and demonstrated the unique opportunities for both clinical utilities and biological insights provided by these representations, or latent factors. We performed an in-depth investigation on the latent factors with deep phenome data of the UK Biobank. First, we explored the relationship between the ECG latent factors and CMR by studying the participants who had both 12-lead ECG and CMR acquired at the same visit. We showed that both cardiac structures and functions estimated from the CMR, which is a more resource-intensive and less available modality, could be inferred from latent factors of the readily accessible ECG. Additionally, not only the latent factors were associated with risk of a wide range of cardiometabolic diseases in a cross-sectional analysis but were also predictive of CVD mortality in a median 3.4 years of follow-up.

We further explored the genetics of the latent factors by GWAS followed by a series of bioinformatic analyses to functionally characterize the genetic signals. We identified a total of 170 genetic loci reaching the genome wide significance (p < 5× 10^-8^). The replication of many loci already associated with classical ECG descriptors demonstrated the utility of the deep representation learning in capturing biologically relevant information. More importantly, we identified 29 novel loci in the current study with our latent factors in around 40,000 participants. The number of genetic loci discovered in current study was on par with recent large scale meta-analyses on the individual ECG fragments with >250,000 individuals^2,3^. Of note, a substantial proportion of the genetic signals including 13 novel loci were discovered through the latent factors with subtle morphology (small phenotypic variance). Our findings highlighted the strength of deep learning in harnessing information from high dimensional data such as complete ECG which might otherwise be omitted in the segmented ECG parameters.

Tissue expression analyses of the genetic signals showed strong enrichment in the heart, muscle and vascular tissues, corroborating the most prominent genetic loci identified in the GWAS. For instance, we identified *SCN5A*, *KCND3*, *KCNQ1* and *KCNE1*, genes with well-established associations to various arrhythmic disorders^30,31^ along with cardiomyopathy genes including *DPT*, *TTN*, *PLN* (at the *SLC35F1* locus), *BAG3* and *MYH7B* ^32^. Gene-set analyses revealed that the genetic signals could be parcellated into interlinked aspects of cardiac physiology namely, development, contractility and electrophysiology. Considering the novel, recent studies reported *CDH18* to be a cardiac development gene specifically expressed in the fetal epicardium^33^; *ERBB4*, *MECOM*, *GPR126* and *NRP1* have been implicated in cardiovascular development. The actin filament-associated protein 1 and kalirin, encoded by *AFAP1* and *KALRN* respectively, are mechanosensing proteins^34–42^. *AKIP1* in the *ST5* locus has been found to promote cardiomyocyte hypertrophy in response to stress^43,44^. We identified *DPP6*, a gene implicated in familial form idiopathic ventricular fibrillation in Dutch population and was associated with QT interval albeit at a suggestive level (*P*=1.66×10^-6^) in the GWAS of African Americans ^45,46^. *DPP6* encodes an auxiliary subunit that modulates the voltage-gated potassium channel Kv4.3 responsible for transient outward potassium current (I_to_) in the heart^47^.

Our finding also demonstrated the uncoupling of the genetic and environmental components of the ECG. First, the latent factors are different in heritabilities indicating the different genetic loadings. For example, both factor 10 and factor 30 captured phenotypic variation of the ventricular rate and the QT interval but genetic loading was higher for factor 30 which was reflected in higher SNP heritability as well as higher number of genetic loci identified. Association patterns with the cardiometabolic diseases were also not identical between the phenotypic association and genetic correlation analysis. For example, factor 21, factor 25 and factor 28 showed concordant associations with bundle branch block in both analyses but factor 18 was only correlated with the disease on the genetic level. Similarly, factor 4 and factor 30 showed moderate genetic correlation with heart failure despite non-significant phenotypic associations. We further explored this uncoupling by visualizing the effects of genetic variants on the ECG by leveraging the decoder of the VAE model. We additionally made both the decoder (https://genetics.ecgx.ai/) and encoders (https://encoder.ecgx.ai) available online to enable further exploration of current results as well as analysis of new data.

Our findings are subject to several limitations. First, we modelled a high-dimensional electrocardiogram with a latent space of 32 dimension using the β-VAE which was optimized iteratively. We noted that higher dimensions could yield a better reconstruction but also at the potential cost of interpretability and model’s ability to interpolate. Capturing rare, subtle but clinically relevant differences in the ECG became a bigger challenge in a relatively healthy cohort such as the UK Biobank^48^. Finetuning of the model would be required to adapt the latent space to a new cohort from a different population. It is also important to point out that statistical independence between the latent factors did not reflect separation of biological functions. On the contrary, our analyses showed that the overlaps in biology, albeit in a different proportion, were captured by the latent factors. The cardiac parameters estimated from CMR were produced automatically by a validated DL-based pipeline which included a QC step to remove erroneous results; direct inspections were not performed. The participants in the UK Biobank are primarily Europeans which may limit the applicability of our findings in other ethnic groups. We have prioritized the candidate causal variants and genes through the bioinformatic analyses only. Further experimental validations of these candidates are needed.

In summary, we applied representation learning on full 12-lead ECG into 32 latent factors using a generative deep learning model. We showed these latent factors are interpretable and revealed novel insights.

## Data Availability

Programming code to train and use the FactorECG model is available through: https://github.com/rutgervandeleur/ecgxai. An online tool to convert any ECG into its FactorECG is available through: https://encoder.ecgx.ai. UK Biobank data is available upon application through the UKB Showcase https://www.ukbiobank.ac.uk.

## Acknowledgments

This research has been conducted using the UK Biobank resource under application numbers 74395. We thank all UK Biobank participants for their contributions. We thank Bjarne de Jong for his contribution to the processing the ECG data. We thank the Center for Information Technology of the University of Groningen for their support and for providing access to the Peregrine high-performance computing cluster. In addition, we thank the “Medische en Informatie Technologie Systeembeheer” of the University Medical Center of Groningen for their support and maintenance of our own computing cluster.

## Code Availability Statement

Programming code to train and use the FactorECG model is available through: https://github.com/rutgervandeleur/ecgxai. An online tool to convert any ECG into its FactorECG is available through: https://encoder.ecgx.ai.

