## Supplementary_figures for "Deep representation learning of electrocardiogram reveals novel insights in cardiac structure and functions and connections to cardiovascular diseases"

**Contents:**

**Figures S1-S7**

**Figure S1**

**Distribution of the latent factors by age and sex.**

2D kernel density plots of all latent factors stratified by age (y-axis) in sex subgroups. Purple: women; Green: men.

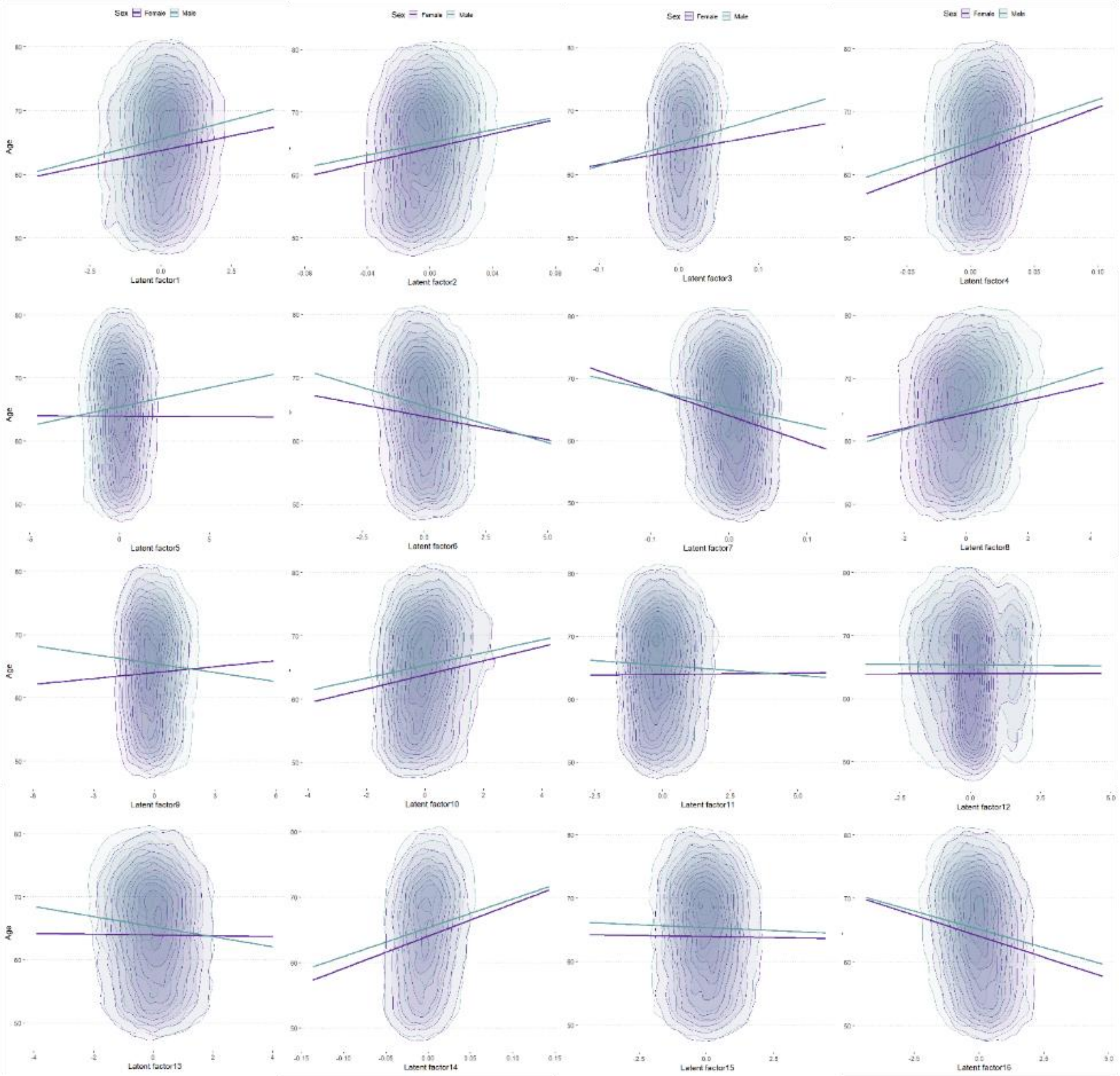

**Figure S1 - continued**

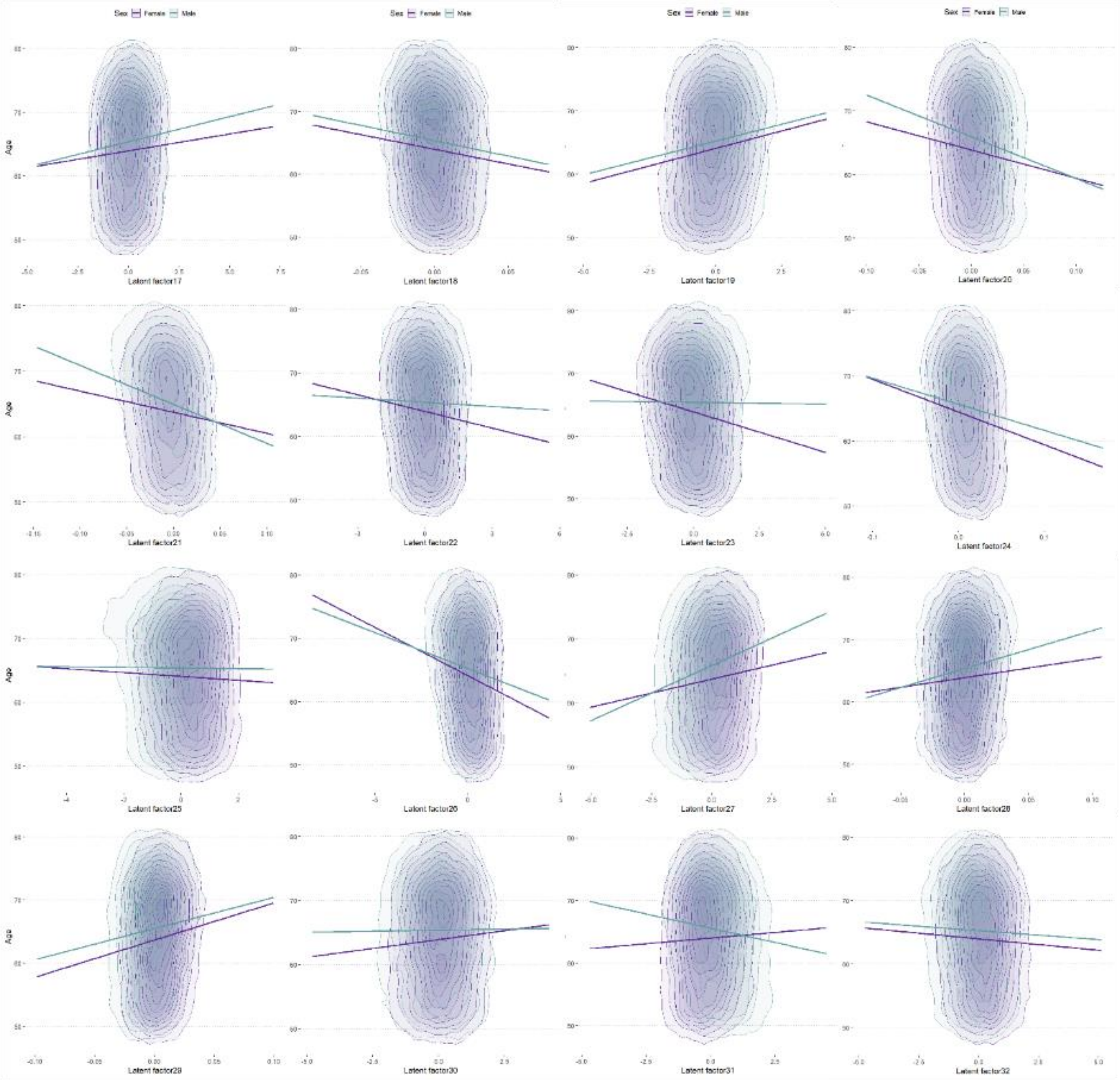

#### Figure S2 Factor traversals for visualizing the effect of individual ECG factors on the ECG morphology

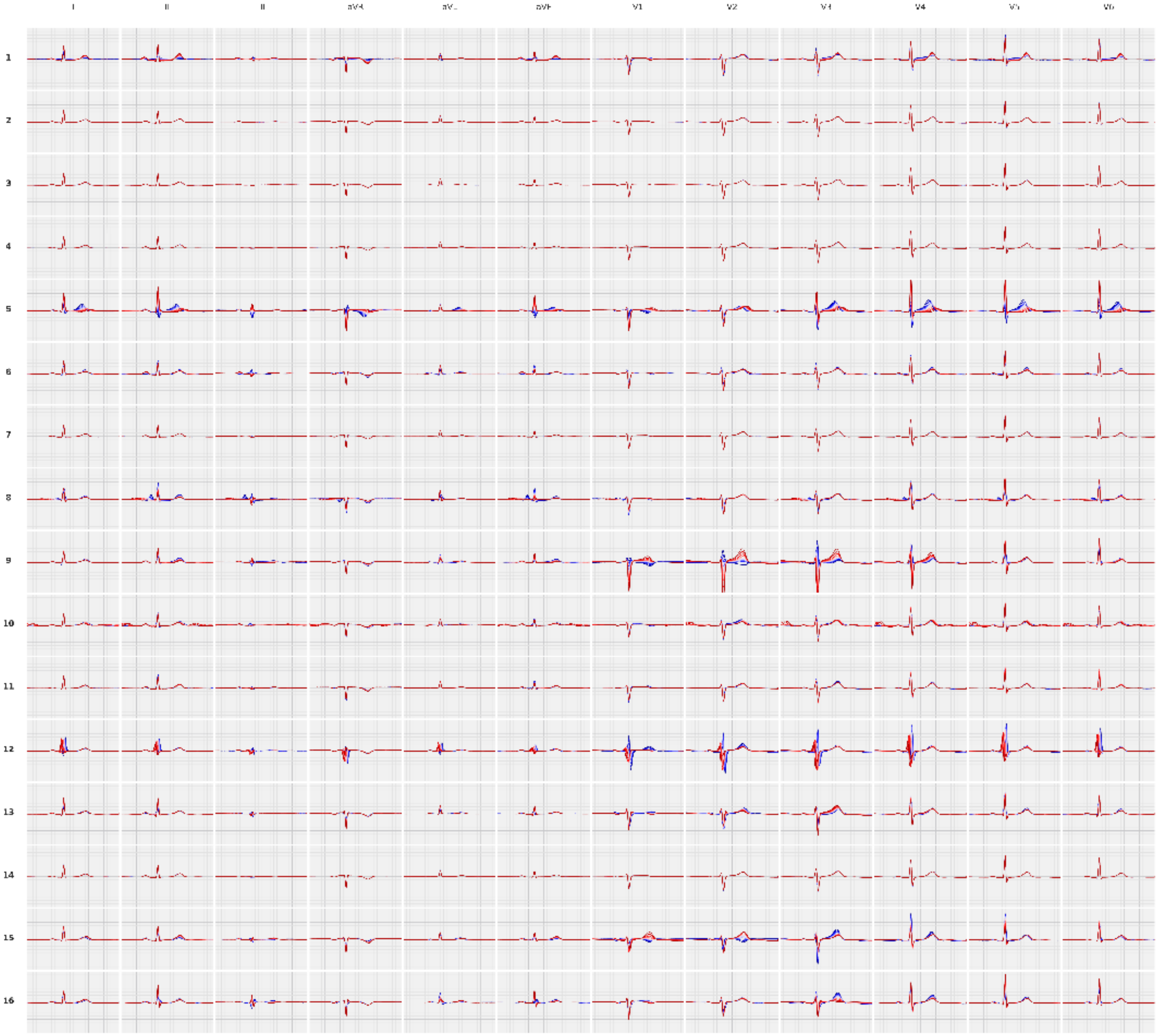

### Figure S2 - continued

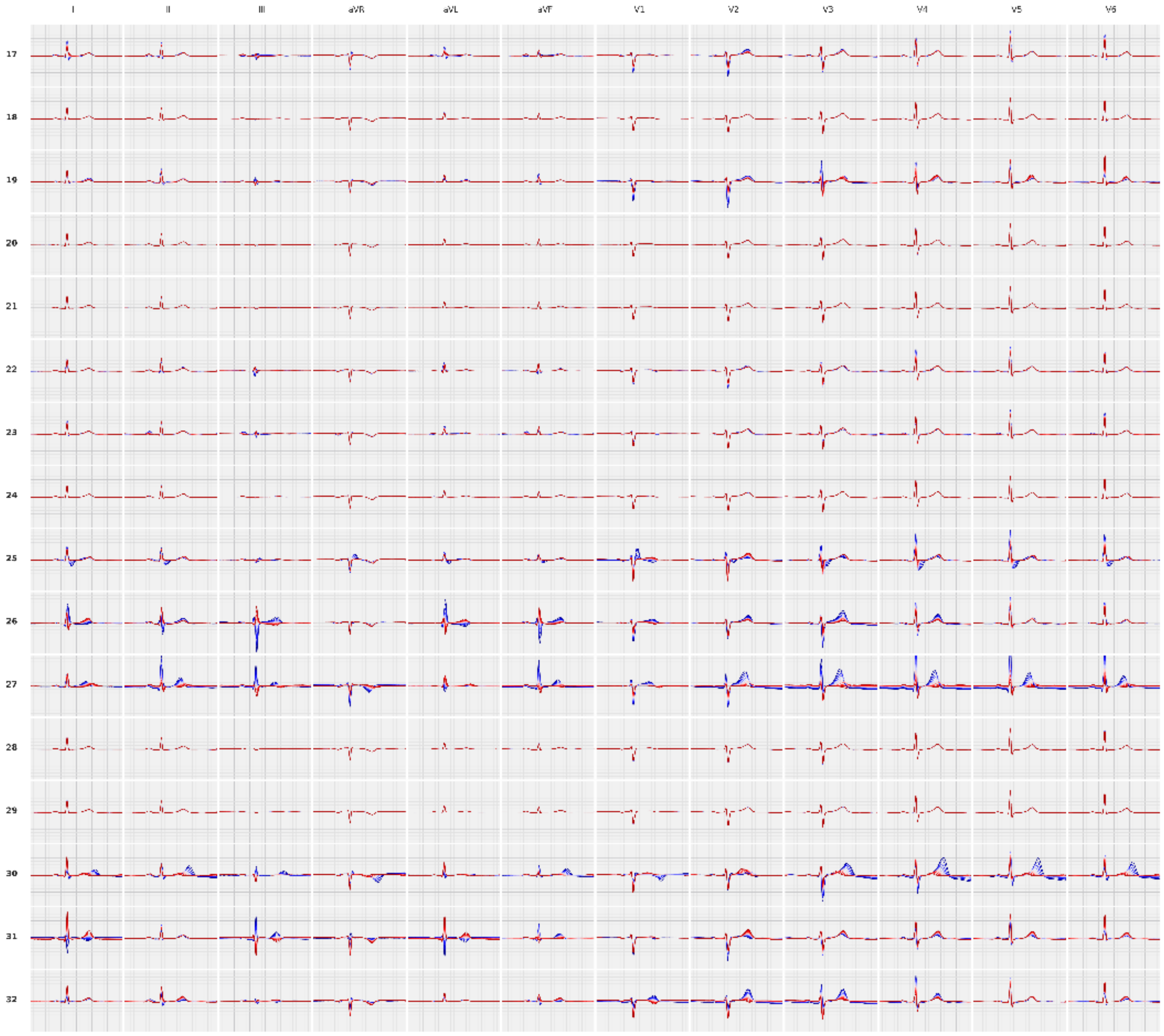

**Figure S3 Phenotypic and genetic correlations between the latent factors.**

Top right: Pearson correlations between the latent factors; Bottom left: genetic correlations estimated using LD-score regression for latent factors with GWAS summary statistics passing quality control.

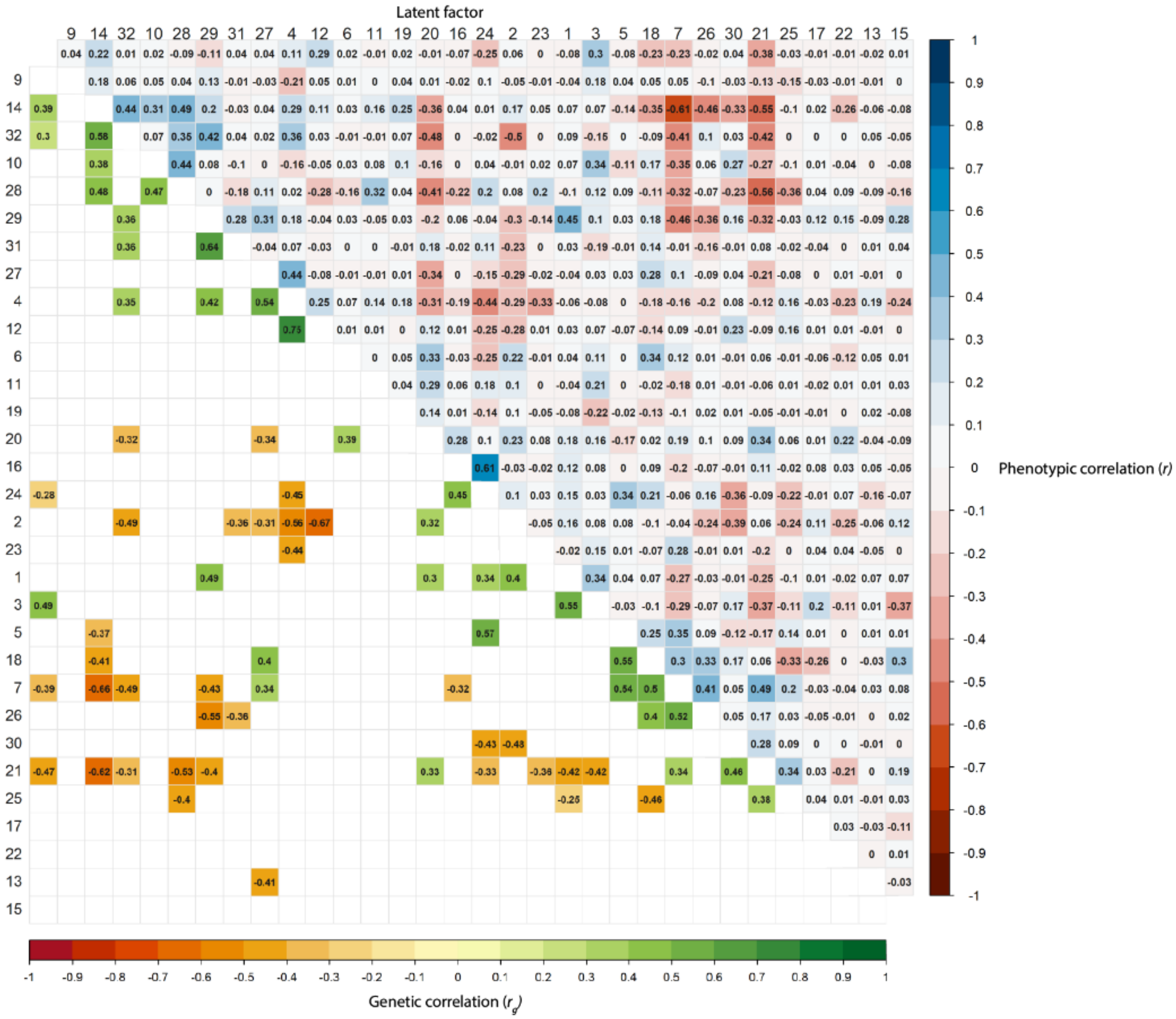

**Figure S4 Forest plot for cardiovascular mortality predicted by latent factors**

Cardiovascular mortality prediction by the latent factors in cox regressions adjusting for age and sex. Hazard ratio (HR) per standard deviation change of the latent factor is shown.

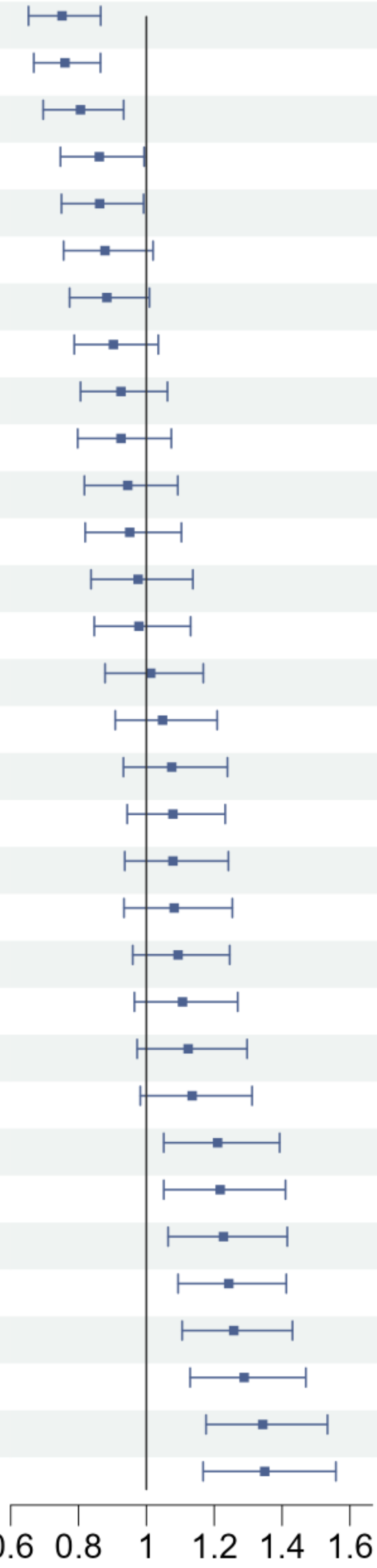


| Predictor | HR (95% CI) | p-value |
|---|---|---|
| latent factor 20 | 0.75 (0.65 - 0.87) | 7.2e-05 |
| latent factor 21 | 0.76 (0.67 - 0.86) | 3.1e-05 |
| latent factor 1 | 0.81 (0.70 - 0.93) | 0.0039 |
| latent factor 24 | 0.86 (0.75 - 0.99) | 0.04 |
| latent factor 22 | 0.86 (0.75 - 0.99) | 0.038 |
| latent factor 31 | 0.88 (0.76 - 1.02) | 0.088 |
| latent factor 26 | 0.88 (0.77 - 1.01) | 0.067 |
| latent factor 7 | 0.90 (0.79 - 1.04) | 0.14 |
| latent factor 25 | 0.93 (0.81 - 1.06) | 0.27 |
| latent factor 16 | 0.93 (0.80 - 1.07) | 0.31 |
| latent factor 19 | 0.95 (0.82 - 1.09) | 0.45 |
| latent factor 2 | 0.95 (0.82 - 1.10) | 0.51 |
| latent factor 15 | 0.98 (0.84 - 1.14) | 0.75 |
| latent factor 17 | 0.98 (0.85 - 1.13) | 0.76 |
| latent factor 29 | 1.01 (0.88 - 1.17) | 0.86 |
| latent factor 6 | 1.05 (0.91 - 1.21) | 0.52 |
| latent factor 18 | 1.07 (0.93 - 1.24) | 0.32 |
| latent factor 11 | 1.08 (0.94 - 1.23) | 0.27 |
| latent factor 23 | 1.08 (0.94 - 1.24) | 0.3 |
| latent factor 4 | 1.08 (0.93 - 1.25) | 0.29 |
| latent factor 12 | 1.09 (0.96 - 1.25) | 0.18 |
| latent factor 5 | 1.11 (0.96 - 1.27) | 0.15 |
| latent factor 32 | 1.12 (0.97 - 1.30) | 0.11 |
| latent factor 30 | 1.13 (0.98 - 1.31) | 0.086 |
| latent factor 3 | 1.21 (1.05 - 1.39) | 0.0079 |
| latent factor 27 | 1.22 (1.05 - 1.41) | 0.0086 |
| latent factor 13 | 1.23 (1.06 - 1.42) | 0.0049 |
| latent factor 9 | 1.24 (1.09 - 1.41) | 0.00087 |
| latent factor 28 | 1.26 (1.11 - 1.43) | 0.0005 |
| latent factor 14 | 1.29 (1.13 - 1.47) | 0.00017 |
| latent factor 10 | 1.34 (1.18 - 1.53) | 1.4e-05 |
| latent factor 8 | 1.35 (1.17 - 1.56) | 4.9e-05 |

**Figure S5 Forest plot for cardiovascular mortality predicted by traditional ECG parameters**

Cardiovascular mortality prediction by the traditional ECG parameters in cox regressions adjusting for age and sex. Hazard ratio (HR) per standard deviation change of the traditional ECG parameters is shown.

| Predictor | HR (95% CI) | p-value |
|---|---|---|
| R axis | 0.90 (0.78 - 1.04) | 0.15 |
| P axis | 0.92 (0.80 - 1.07) | 0.28 |
| QT interval | 0.98 (0.85 - 1.12) | 0.74 |
| PR interval | 1.14 (1.00 - 1.29) | 0.049 |
| T axis | 1.15 (1.02 - 1.30) | 0.027 |
| QRS duration | 1.22 (1.10 - 1.36) | 0.00027 |
| QTc interval | 1.25 (1.14 - 1.36) | 7.3e-07 |
| Ventricular rate | 1.28 (1.13 - 1.45) | 7.1e-05 |

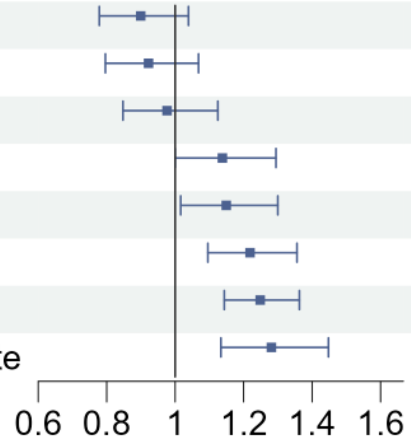

**Figure S5 Regional plots of the novel loci identified in the GWAS.**

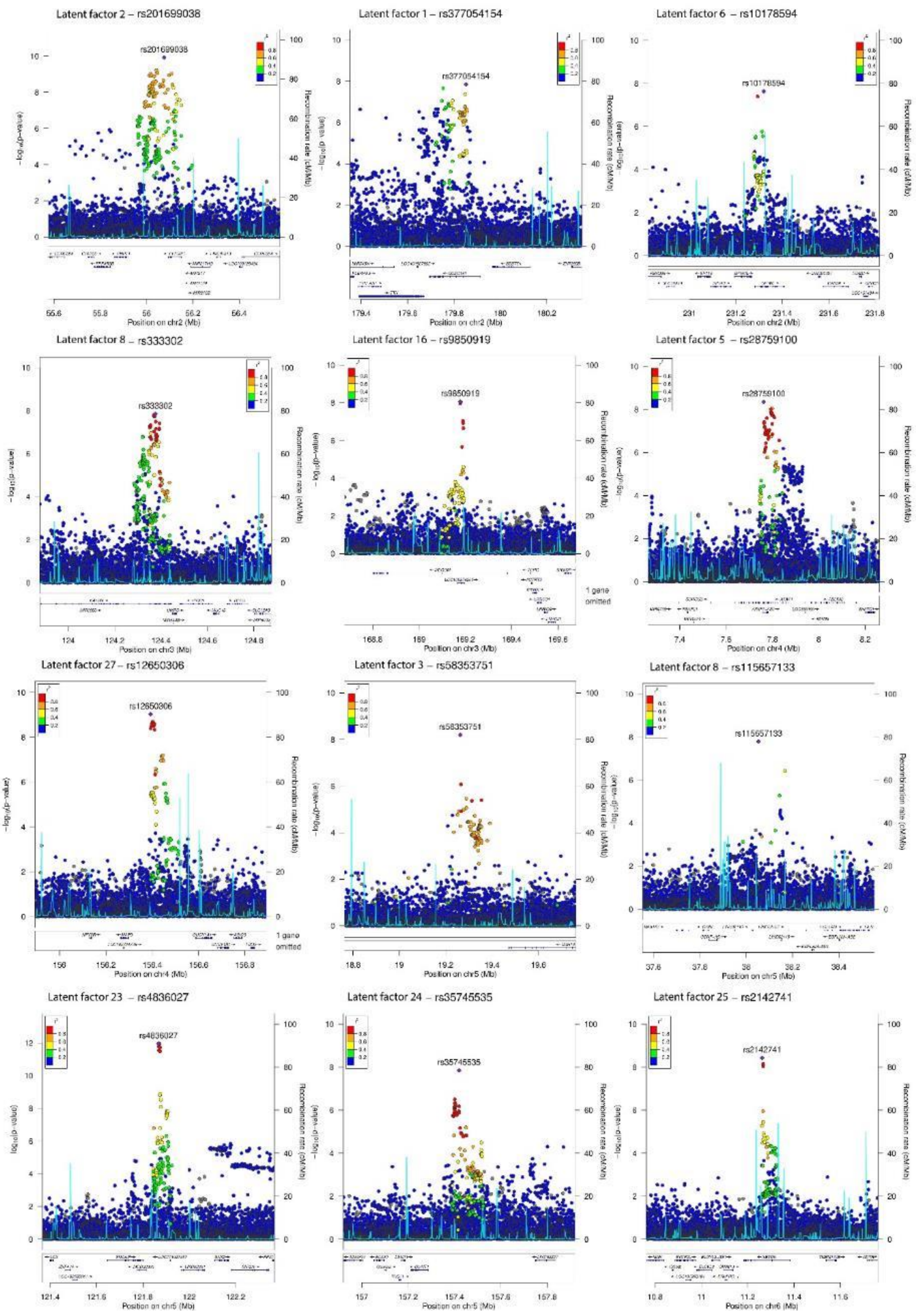

**Figure S5 – continued**

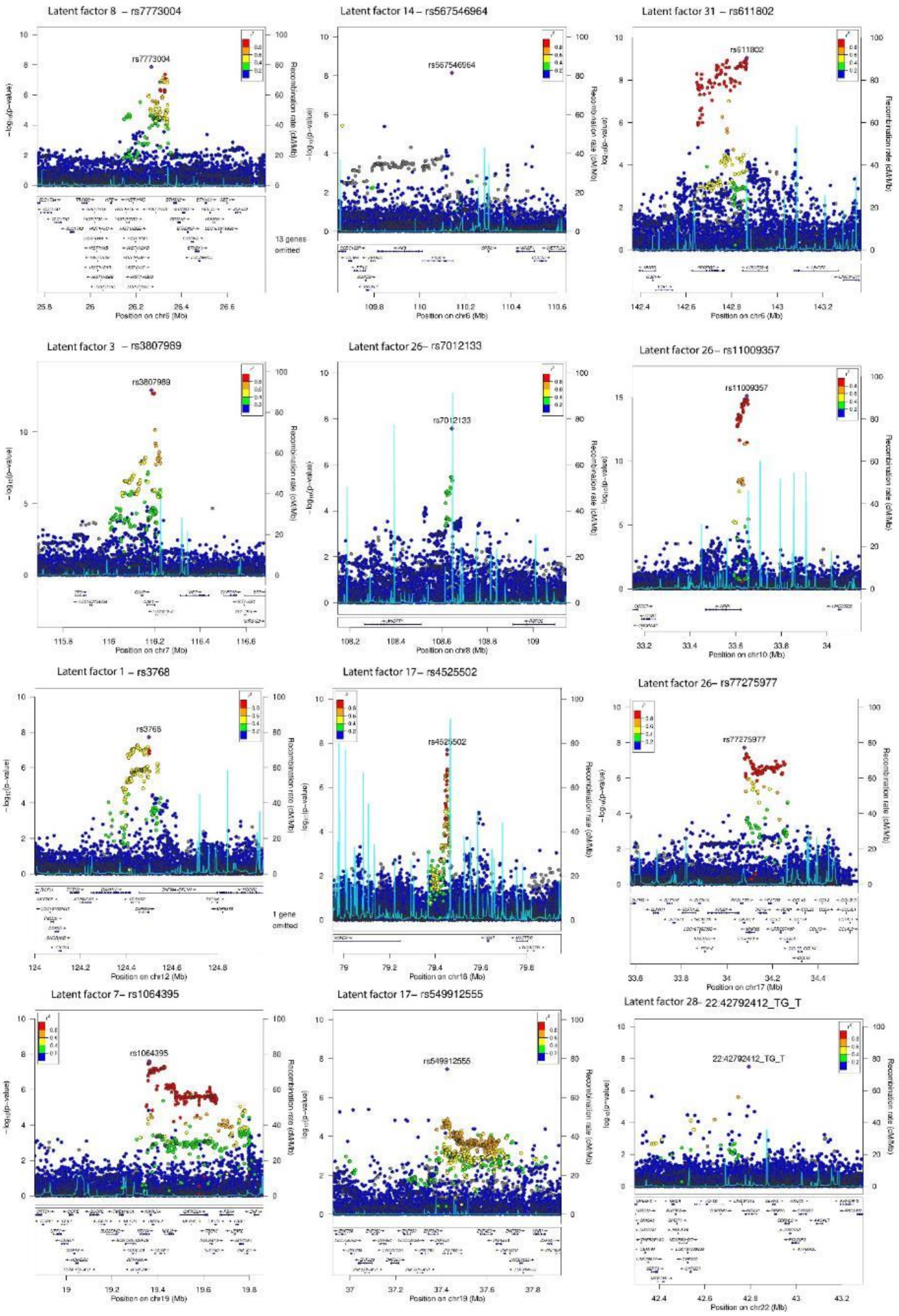

**Figure S6 Interactive application for latent factors on ECG morphology**

The effect per allele on each dimension for decoded back to the VAE model to visualize the changes in ECG morphology. The app can be accessed at [https://genetics.ecgx.ai/](https://genetics.ecgx.ai/).

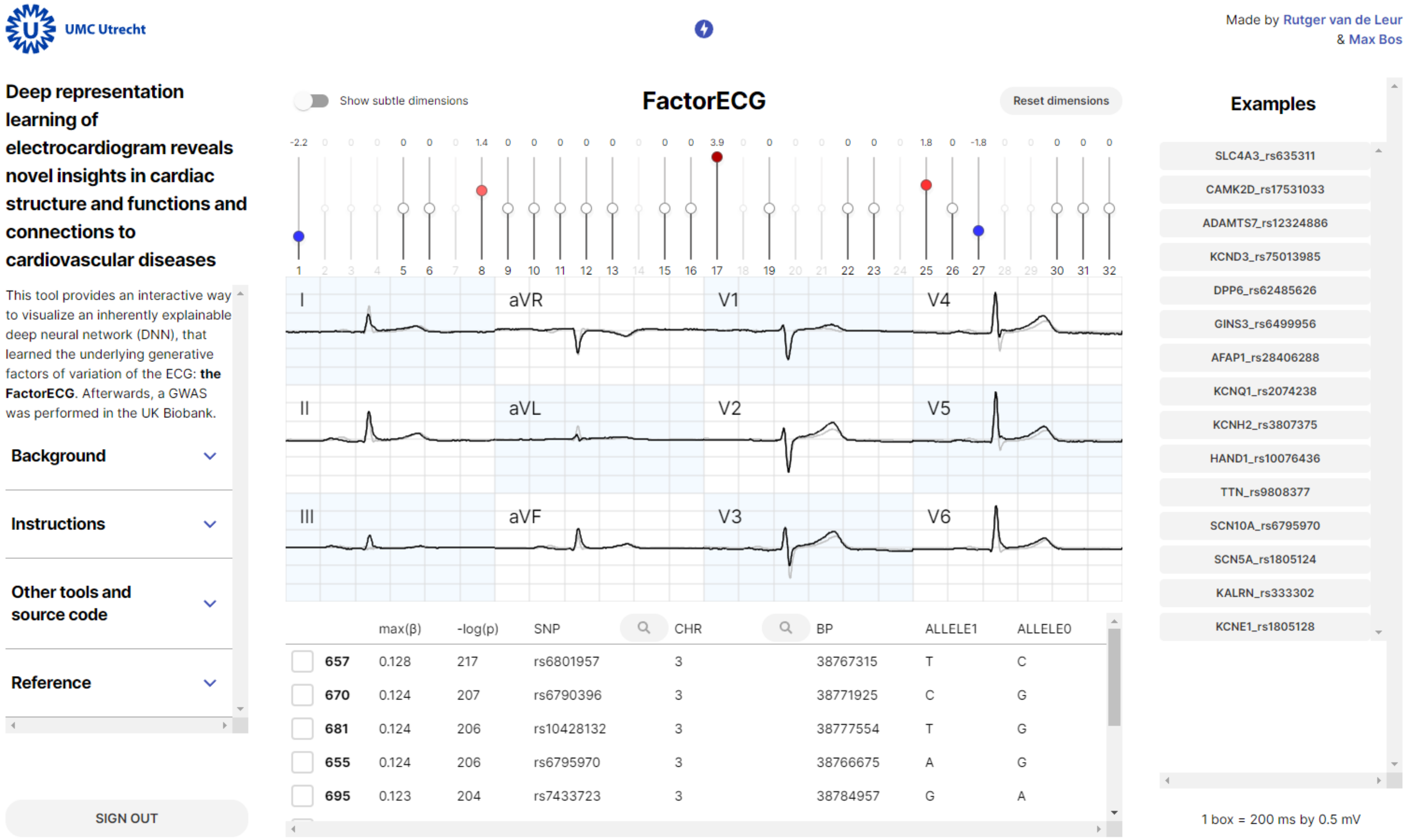

**Figure S7 Results from ontology term clustering by semantic similarity**

Semantic similarities between the Gene Ontology (GO) associated gene-set by three GO categories, biological processes (BP) , molecula fucntions (MF) and cellular components (CC). Clustering was performed using the binary cut algorithm and results annotated by word cloud of shared words among the gene-sets.  Font size reflects the enrichment of the keywords in each cloud.

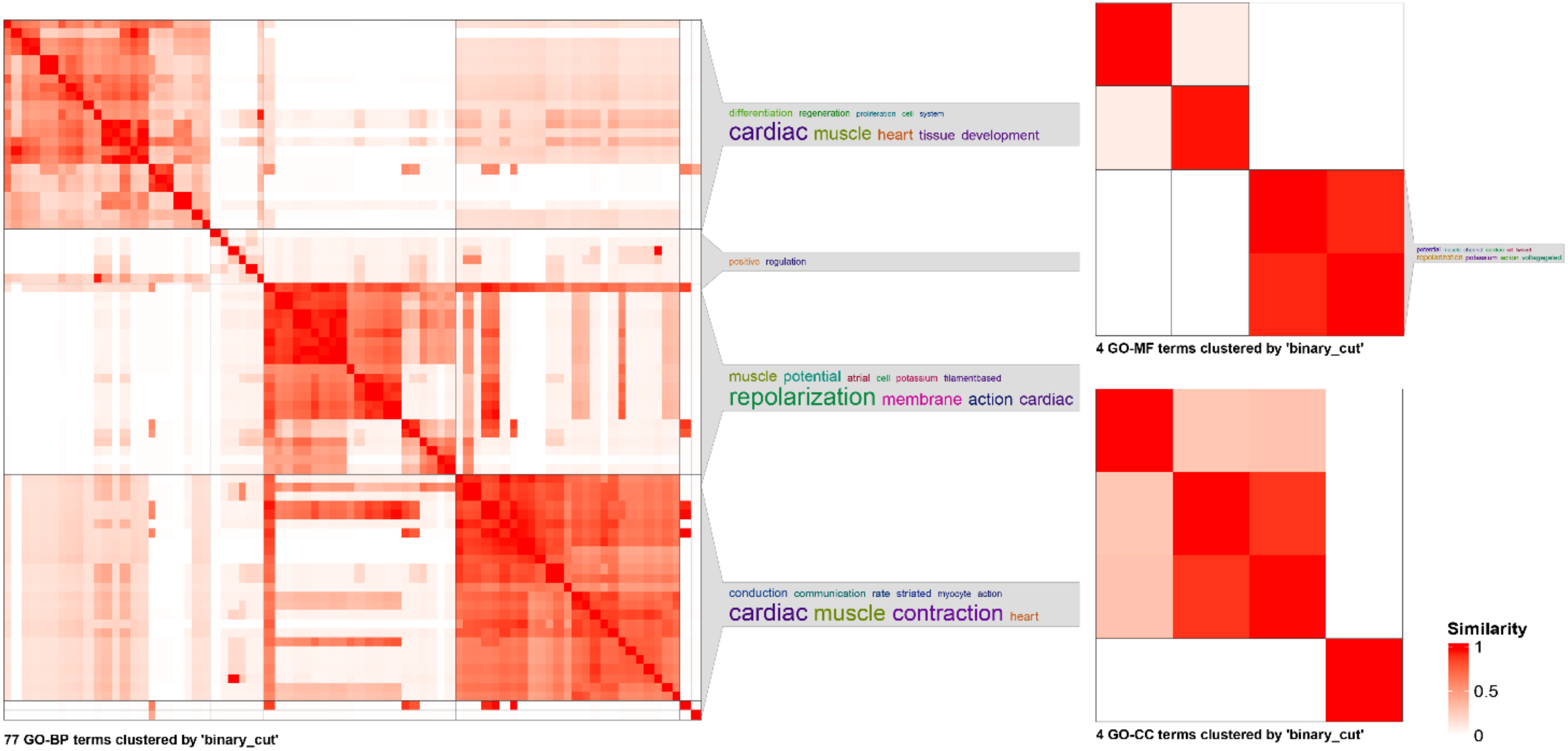

**Figure S8 Genetic correlation of latent factors with cardiometabolic diseases.**

Dot plot shows the genetic correlation of the latent factors as a risk factor for selected cardiometabolic disease. The colour of the dot indicates the coefficients in genetic correlations estimated with LD score regression. Coefficients were adjusted for age and sex. The size of dot reflects the p-value categories for the latent factor in the corresponding logistic regression model; $p=8.22\times10^{-5}$ is the Bonferroni corrected p-value taking $\alpha=0.05$. The bar plot shows the SNP heritability of the disease or latent factor.

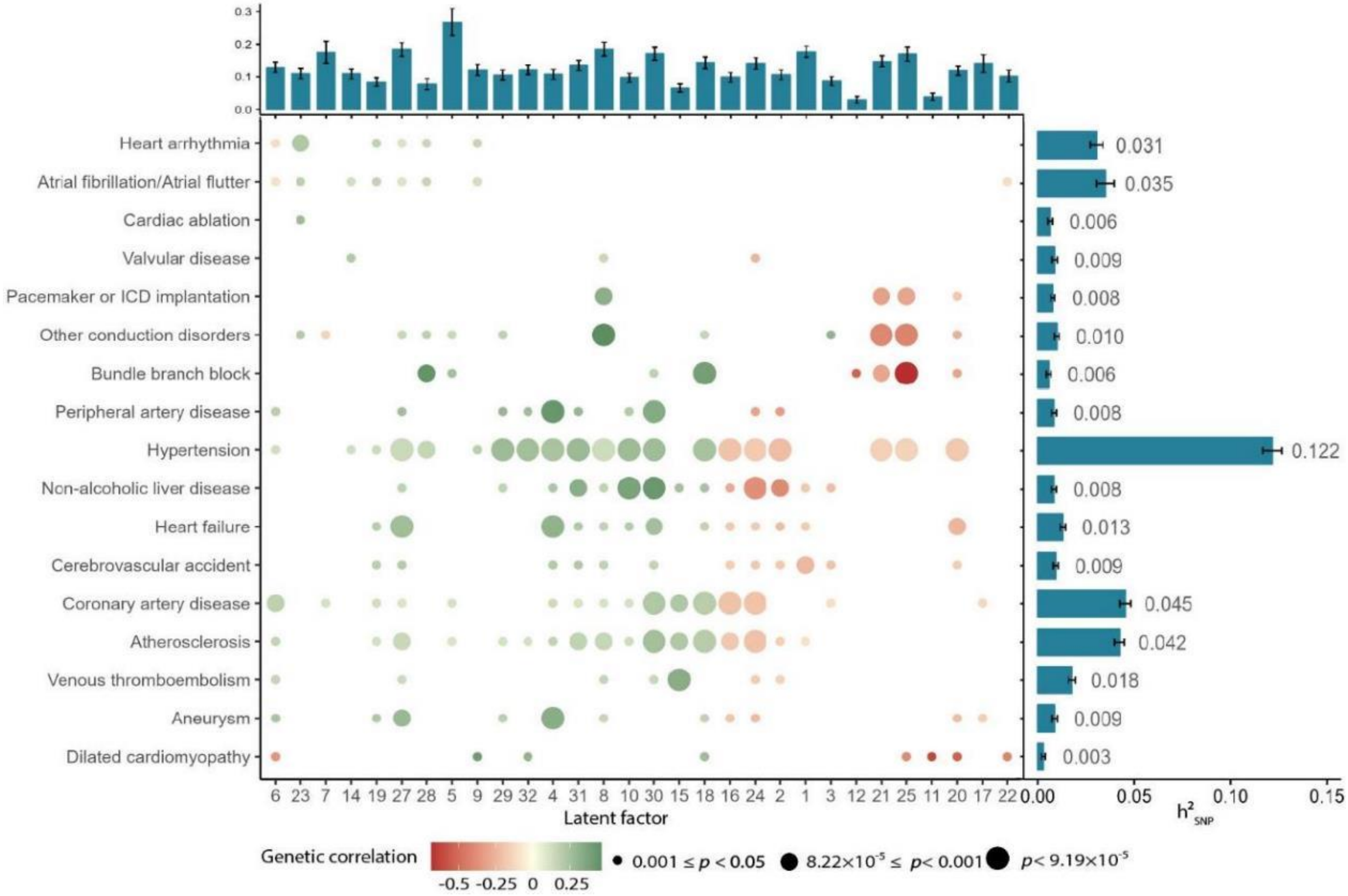

**Figure S9 Trait-locus associations reported in GWAS catalog for novel loci.**

For each new locus associated with ECG morphology, the lead variant, and all variants in LD>0.8 were queried against the GWAS catalog to identify associations reported in previous GWAS. Each dot represents one association record in the GWAS catalog, coloured by curated biological functional groups.

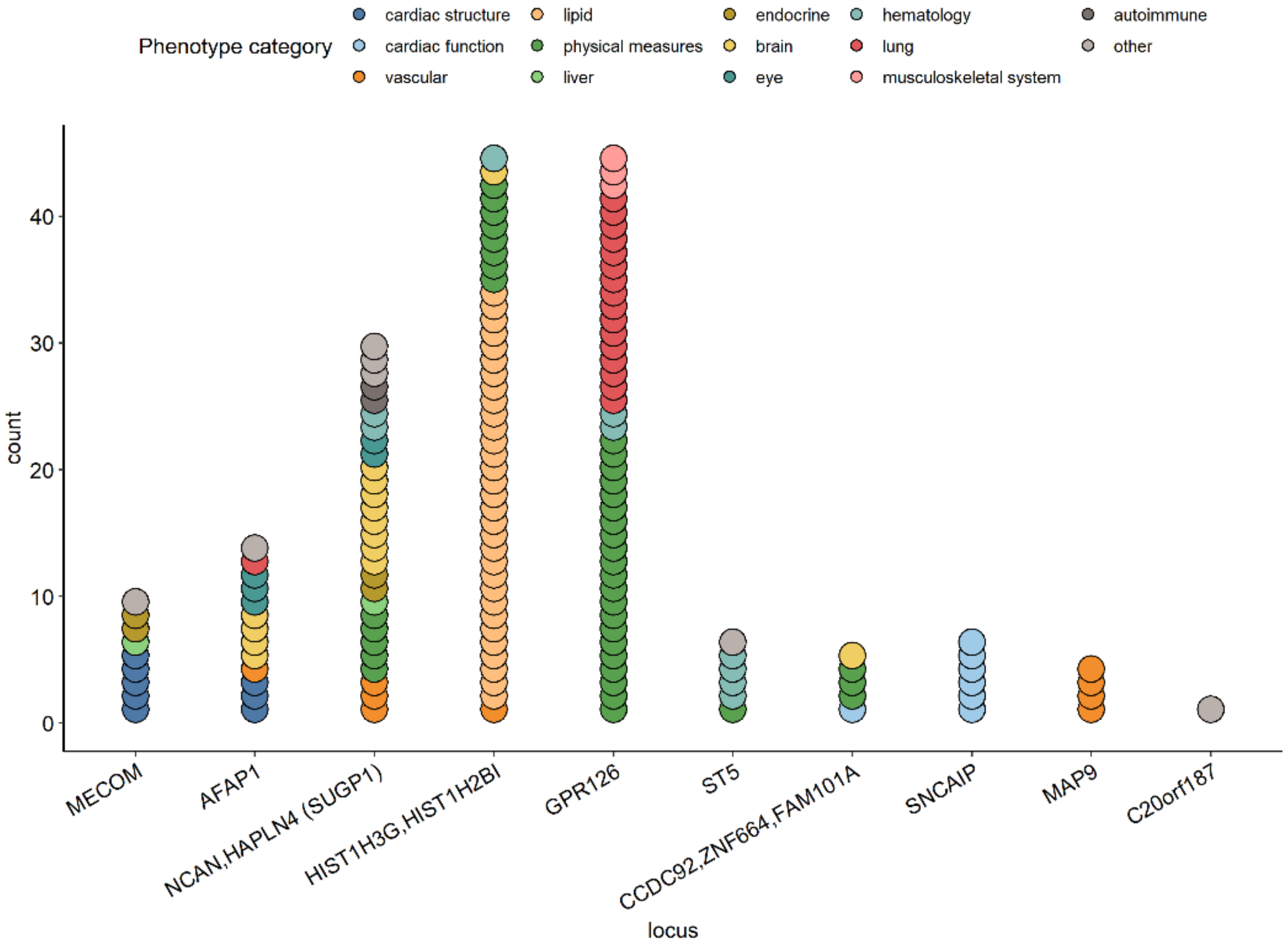
